# Detection of clonal hematopoiesis of indeterminate potential via genome or exome sequencing profoundly underestimates disease associations

**DOI:** 10.1101/2025.08.11.25333294

**Authors:** Robert Corty, Yash Pershad, Caitlyn Vlasschaert, Leo Luo, Taralynn Mack, Kaushik Amancherla, Cassianne Robinson-Cohen, Michael Savona, Alexander G. Bick

**Author notes:** **Corresponding author:** Alexander Bick, 550 Robinson Research Building, 2200 Pierce Ave, Nashville, TN 37232. RWC and YP (second listed author) are co–first authors. The authorship order reflects that the study was initiated by RWC, who was joined by YP (second listed author) in leading the project.

## Abstract

Clonal hematopoiesis of indeterminate potential (CHIP) occurs when at least 4% of blood cells harbor somatic mutations in leukemogenic genes and is associated with increased risk for cardiovascular disease, malignancy, and mortality. While deep sequencing (>1,000x coverage) is the gold standard for CHIP detection, most large-scale studies rely on shallow genome/exome sequencing (∼35x coverage) from biobanks. However, the sensitivity and specificity of genome-based CHIP detection remain unknown, raising concerns about the accuracy of reported disease associations. We performed both deep targeted sequencing and genome sequencing on identical DNA samples from 9,925 participants to characterize genome-based CHIP detection performance. Genome sequencing showed poor sensitivity (28%) and positive predictive value (44%) compared to deep sequencing, with performance highly dependent on clone size. Simulation studies revealed that these ascertainment errors dramatically reduce statistical power and underestimate true effect sizes by >80%. These findings indicate that genome-based studies profoundly underestimate CHIP-disease associations, necessitating targeted deep sequencing for accurate clinical risk assessment.

## Main

Clonal hematopoiesis of indeterminate potential (CHIP) occurs when ≥4% of nucleated blood cells harbor a somatic mutation in a leukemogenic gene (1). The gold standard method to detect CHIP is deep (>1,000x) sequencing of a peripheral blood (2, 3). Researchers have also detected CHIP using shallow (∼35x) sequencing of genomes or exomes in large biobanks (4). They found that CHIP is associated with heightened risk for a wide range of diseases including cardiovascular disease, solid-organ malignancy, auto-immune disease, kidney disease, and all-cause mortality (5, 6).

A critical gap in the field is that the sensitivity and specificity of genome-sequencing-based CHIP detection are not known, nor are the effects of CHIP ascertainment errors on epidemiologic associations. Errors in CHIP ascertainment could render the estimates of CHIP-attributable risk inaccurate. Accurate estimation of the CHIP-attributable risk for disease is critical for clinical applications such as risk stratification, disease monitoring, and treatment selection.

To address this gap, we performed both genome sequencing and deep sequencing on the same DNA sample from 9,925 research participants and characterized the testing properties of genome-sequencing-based CHIP detection. Using the empiric sensitivity and specificity of genome-sequencing-based CHIP detection, we simulated how ascertainment error influences the power and precision of CHIP-disease association studies. For deep sequencing, we performed error-corrected targeted sequencing of the exons of CHIP driver genes with median depth after deduplication of ∼1700x (2). Genome sequencing was performed using the Illumina DNA PCR-Free Prep targeting a median depth of 30x. To detect CHIP, *Mutect2* was used to call somatic mutations in CHIP driver regions. We filtered variants based on read depth (≥100 for deep sequencing, ≥ 15 for genome sequencing), variant allele read depth (≥3 for deep sequencing, ≥ 2 for genome sequencing), double-strand support, and inconsistency with germline heterozygosity, and variant allele fraction (VAF) ≥ 2% (4).

Among 9,925 participants, we identified 1,255 (13%) with CHIP by genome sequencing and 1,509 (15%) with CHIP mutations by deep sequencing. We calculated the sensitivity and positive predictive value (PPV) of genome-sequencing-based CHIP calling on a per-variant level using deep-sequencing-based calls as the gold-standard and genome-sequencing-based calls as the index test. Genome-sequencing-based CHIP calling had a sensitivity of 28% (426/1,509), specificity of 94% (8,258/8,794), and PPV of 44% (426/962). These performance metrics were highly dependent on the size of the CHIP clone, as quantified by VAF. Sensitivity was 9% for VAF 2-5%, 32% for VAF 5-10%, 67% for VAF 10-20%, and 86% for VAF > 20% (**Figure 1A**). PPV was 0%, 26%, 44%, and 58% across the same VAF bins (**Figure 1B**).

**Figure 1.**
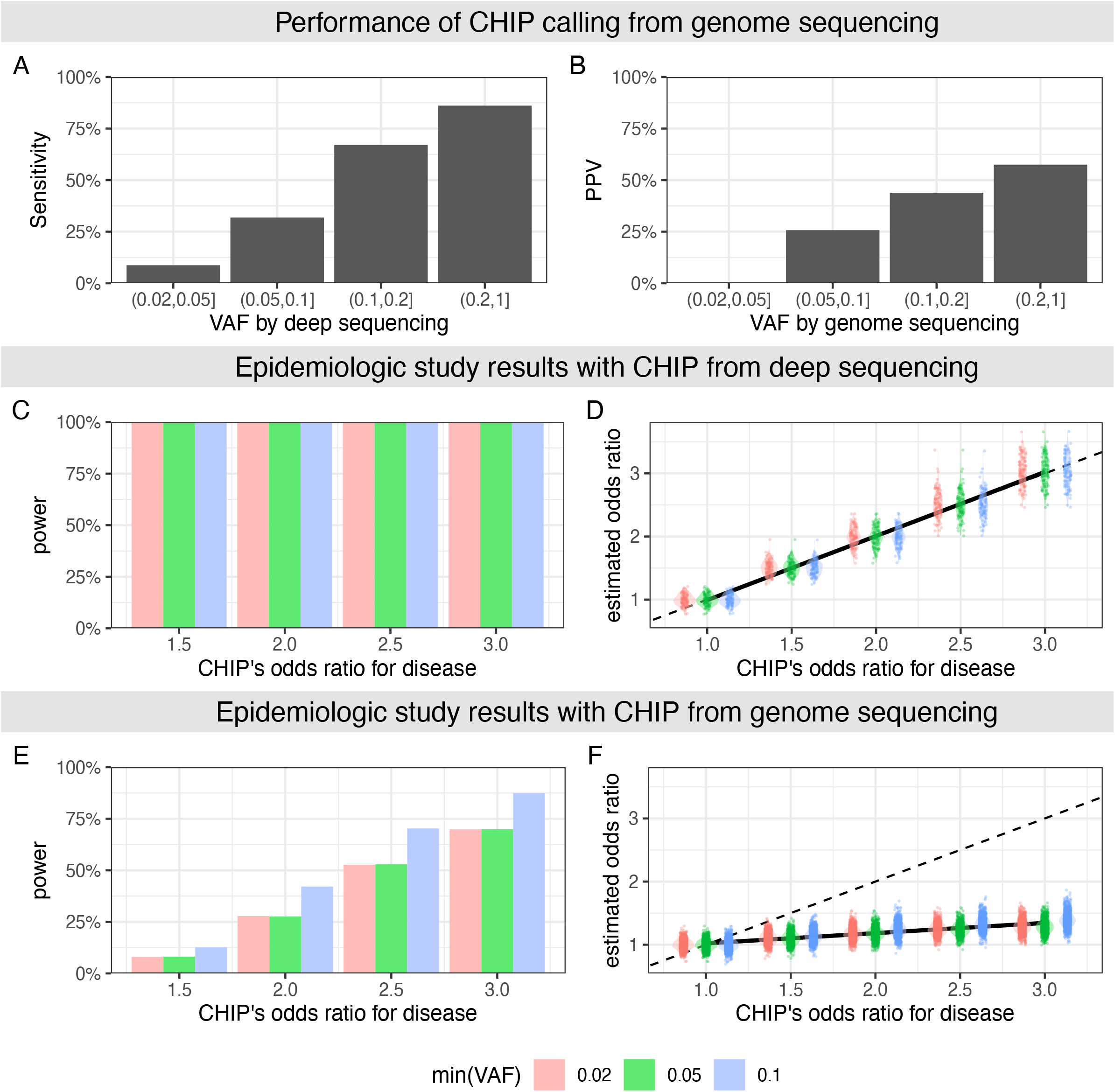
Performance characteristics of genome-sequencing-based CHIP calling and impact on epidemiologic associations. Empiric performance of genome-sequencing-based CHIP detection compared to the gold standard, deep sequencing, stratified by VAF for (A) sensitivity and (B) positive predictive value. Simulated CHIP-disease associations using logistic regression across odds ratios, with minimum VAF thresholds of 0.02, 0.05, and 0.1 using deep-sequencing-based CHIP detection for (C) statistical power and (D) odds ratio estimation compared to true odds ratio. As above, simulated CHIP-disease associations using logistic regression, but instead using genome-sequencing-based CHIP detection for (E) statistical power and (F) bias in odds ratio estimation compared to true odds ratio.

We performed simulations to determine how ascertainment errors influence the power and precision of CHIP associations with prevalent and incident disease. For each scenario, we simulated 100,000 persons with random sex, random age (40-79 years), CHIP status and VAF calibrated to age and repeated the simulation 1,000 times.

First, we tested the impact of ascertainment errors in studies of the effect of CHIP on disease prevalence. We simulated disease prevalence based on age, sex, and CHIP status for odds ratios (ORs) ranging from 1.0 to 3.0. Genome-sequencing-based CHIP calls were simulated based on empiric, VAF-dependent sensitivity and specificity reported above. We tested for CHIP-disease association using multivariate logistic regression. With deep-sequencing-based CHIP calls, power was 100% and OR estimation was near perfect (**Figures 1C and Figure 1D**). Using all genome-sequencing-based CHIP calls, the power to detect the association was 5%, 25%, 46%, and 71% for a disease with CHIP-associated OR of 1.5, 2.0, 2.5, and 3.0, respectively. Power was higher when people with genome-sequencing-estimated VAF < 10% were excluded, consistent with the high rate of false positives observed in that group (**Figure 1E**). Using genome-sequencing-based CHIP calls, estimated ORs captured ∼16% of true liability (**Figure 1F**).

Second, we tested the impact of ascertainment errors in studies of the effect of CHIP on disease incidence. We simulated age of disease onset and age of censoring based on age, sex, and CHIP status for hazard ratios (HRs) ranging from 1.0 to 3.0. Genome-sequencing-based CHIP calls were simulated as above. We tested for CHIP-disease association using Cox proportional hazards regression. With deep-sequencing-based CHIP calls, power was 100% and the estimated HR captured 73% of the true HR, consistent with a known downward bias in Cox proportional hazards regression (**Supplementary Figure 3**). Using all genome-sequencing-based CHIP calls, the power to detect the association was 8%, 28%, 53%, and 70% for a disease with CHIP-associated HR of 1.5, 2.0, 2.5, and 3.0, respectively (**Supplementary Figure 3**). As with prevalence, power was higher when people with genome-sequencing-estimated VAF < 10% were excluded. Estimated HRs using genome-sequence-based CHIP calls captured ∼9% of the true hazard for disease (**Supplementary Figure 3**).

Our study has several implications. First, accurate estimation of the strength of association between CHIP and disease necessitates sensitive CHIP detection, which is not possible with genome-sequencing-based CHIP ascertainment. Even though studies of ever-larger biobanks with genome-sequencing-based CHIP calls can overcome deficits in power, ascertainment errors lead to underestimation of the strength of CHIP-disease association regardless of cohort size. Second, since most well-powered CHIP epidemiology studies reanalyze genome or exome sequencing, the widely cited associations between CHIP and disease risk, such as cardiovascular disease, are significantly underestimated. Studies with targeted CHIP sequencing are necessary to understand the true strength of association between CHIP and disease. Third, people with genome-sequencing-based CHIP calls with VAF < 10% contain enough false positives that removing them from the analysis is more beneficial than including them. While it is widely reported in studies that use genome-or exome-based CHIP detection that CHIP with VAF >10% carry a higher risk for many diseases, this observation is likely due to CHIP ascertainment errors.

In summary, errors in CHIP ascertainment in genome and exome-based CHIP calling, lead to (1) insensitive studies, which can be partially rescued by excluding people with observed VAF < 10%, and (2) a profound underestimation of the strength of CHIP-disease association, which can be rescued only with improved CHIP ascertainment – that is, by deep, targeted sequencing.

## Data Availability

A table of CHIP mutations detected by deep targeted sequencing and whole-genome sequencing is available for download on github.com/bicklab/wgs_chip_is_spec_but_not_sens.

## Notes

**Conflict of Interest:** M.R.S. has received honoraria for advisory board membership or consultancy from Bristol Myers Squibb, CTI, Forma, Geron, GlaxoSmithKline/Sierra Oncology, Karyopharm, Ryvu Therapeutics, and Taiho Pharmaceutical; has received research funding from ALX Oncology, Astex Pharmaceuticals, Incyte Corporation, Takeda, and TG Therapeutics; holds equity in Empath Biosciences, Karyopharm, and Ryvu Therapeutics; and has been reimbursed for travel expenses by Astex.

### Competing Interest Statement

M.R.S. has received honoraria for advisory board membership or consultancy from Bristol Myers Squibb, CTI, Forma, Geron, GlaxoSmithKline/Sierra Oncology, Karyopharm, Ryvu Therapeutics, and Taiho Pharmaceutical; has received research funding from ALX Oncology, Astex Pharmaceuticals, Incyte Corporation, Takeda, and TG Therapeutics; holds equity in Empath Biosciences, Karyopharm, and Ryvu Therapeutics; and has been reimbursed for travel expenses by Astex.

### Funding Statement

Vanderbilt University Medical Center BioVU projects are supported by numerous sources: institutional funding, private agencies, and federal grants. These include NIH funded Shared Instrumentation Grants S10OD017985, S10RR025141, and S10OD025092; and CTSA grants UL1TR002243, UL1TR000445, and UL1RR024975. Genomic data are also supported by investigator-led projects that include U01HG004798, R01NS032830, RC2GM092618, P50GM115305, U01HG006378, U19HL065962, R01HD074711.
The sequencing of 250,000 WGS individuals from BioVU, including the 10,310 described here, has been funded by the Alliance for Genomic Discovery consisting of NashBio, Illumina and industry partners Amgen, AbbVie, AstraZeneca, Bayer, BMS, GSK, Merck, and Novo. DNA sequencing was performed at deCODE genetics using Illumina sequencing technology.

### Author Declarations

The Institutional Review Board of Vanderbilt University Medical Center oversees BioVU and gave ethical approval for this work (IRB #201783).

## References

1. Jaiswal S, et al. Age-Related Clonal Hematopoiesis Associated with Adverse Outcomes. N Engl J Med. 2014;371(26):2488–2498.

2. Mack T, et al. Cost-Effective and Scalable Clonal Hematopoiesis Assay Provides Insight into Clonal Dynamics. The Journal of Molecular Diagnostics. 2024;26(7):563–573.

3. Stewart CM, et al. Clonal hematopoiesis detection by simultaneous assessment of peripheral blood mononuclear cells, blood plasma, and saliva. Journal of Clinical Investigation. [published online ahead of print: June 19, 2025]. 10.1172/JCI191256.

4. Vlasschaert C, et al. A practical approach to curate clonal hematopoiesis of indeterminate potential in human genetic datasets. Blood. 2023;blood.2022018825.

5. Jaiswal S, Ebert BL. Clonal hematopoiesis in human aging and disease. Science. 2019;366(6465):eaan4673.

6. Walsh K. The emergence of clonal hematopoiesis as a disease determinant. Journal of Clinical Investigation. 2024;134(19):e180063.

