## Supplemental Material for "Detection of clonal hematopoiesis of indeterminate potential via genome or exome sequencing profoundly underestimates disease associations"

### Supplementary Material

#### Supplemental Methods

*Sex as a biologic variable:*

Sex of the individuals was not considered when defining the sensitivity and positive predictive value of CHIP calls using whole-genome sequencing. In our simulation studies, individuals were assigned random sex which was used as a covariate in logistic regressions and Cox proportional hazard regressions.

*Study participants:*

Our study cohort was comprised of individuals who were sequenced as part of the BioVU Alliance for Genomic Discovery and sequenced using deep sequencing to detect CHIP in cohorts within BioVU as part of four prior studies. Three studies examined CHIP in specific disease states: chronic kidney disease[1], radiation therapy for solid tumors[2], and heart failure requiring transplant[3]. The fourth examined participants who donated multiple samples to study the longitudinal dynamics of CHIP[4].

*Statistics:*

Simulation code is provided at details are provided at github.com/bicklab/wgs_chip_is_spec_but_not_sens. In brief, plausible parameters were used to simulate study cohorts, prevalent disease analysis was conducted with a logistic model, and incident disease analysis was conducted with a Cox proportional hazards model, all in R version 4.3.2.

*Study approval:*

Vanderbilt University Medical Center's Institutional Review Board oversees BioVU and approved this project (IRB #201783).

*Data availability:*

A table of CHIP mutations detected by deep targeted sequencing and whole-genome sequencing is available for download on github.com/bicklab/wgs_chip_is_spec_but_not_sens.

*Code availability:*

Analysis code and simulations are available at: github.com/bicklab/wgs_chip_is_spec_but_not_sens.

### Supplemental Figures

### Supplemental Figure 1:

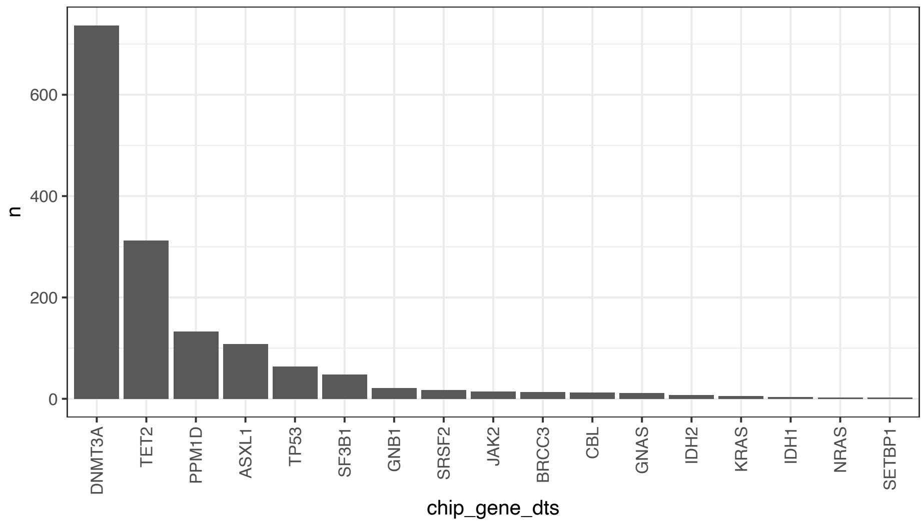


**Supplemental Figure 1. Number of CHIP driver mutations found in each gene by deep, targeted sequencing**

### Supplemental Figure 2:

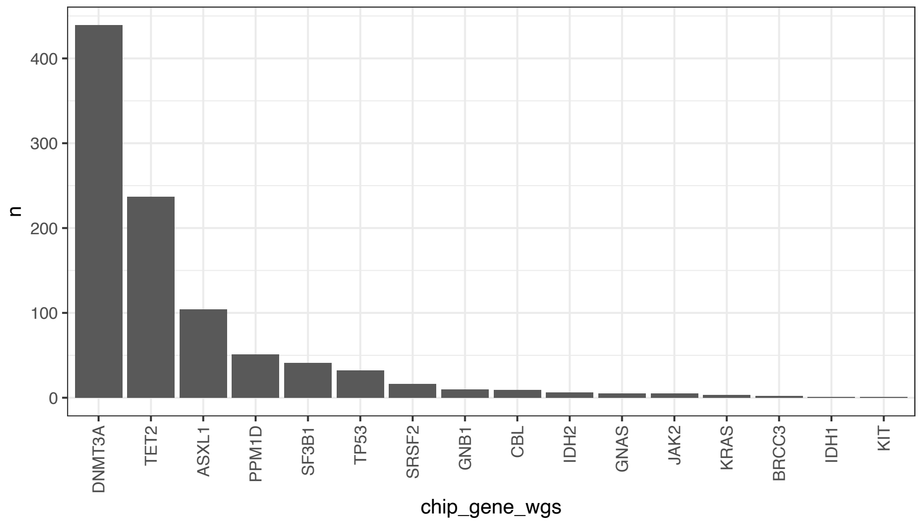


**Supplemental Figure 2. Number of CHIP driver mutations found in each gene by whole genome sequencing (WGS).**

**Supplemental Figure 3:**

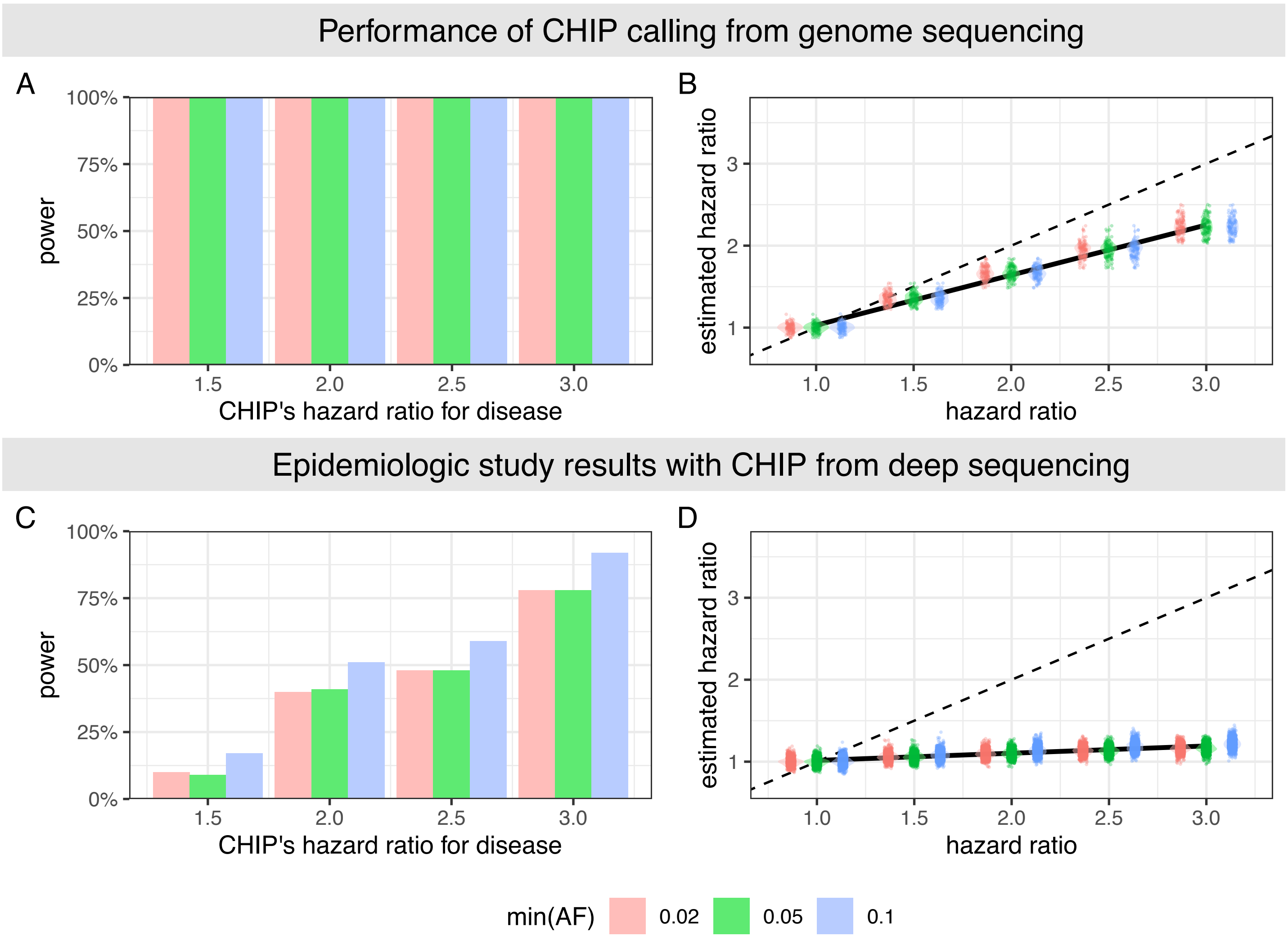


**Supplemental Figure 3. In simulations, incident testing models with access to true CHIP status have 100% power to detect CHIP-disease association and much more precise estimation than models using WGS-based CHIP status.** Simulated CHIP-disease associations using Cox proportional hazards regression across hazard ratios, with minimum VAF thresholds of 0.02, 0.05, and 0.1 using deep-sequencing-based CHIP detection for (C) statistical power and (D) hazard ratio estimation compared to true hazard ratio. As above, simulated CHIP-disease associations using Cox proportional hazards regression, but instead using genome-sequencing-based CHIP detection for (E) statistical power and (F) bias in hazard ratio estimation compared to true hazard ratio. As expected, survival analysis is slightly biased toward the null even with perfect CHIP ascertainment, due to the known random noise injected into the simulation studies, thereby explaining the deviation from y=x.

*Funding and Acknowledgments:*

Vanderbilt University Medical Center's BioVU projects are supported by numerous sources: institutional funding, private agencies, and federal grants. These include NIH funded Shared Instrumentation Grants S10OD017985, S10RR025141, and S10OD025092; and CTSA grants UL1TR002243, UL1TR000445, and UL1RR024975. Genomic data are also supported by investigator-led projects that include U01HG004798, R01NS032830, RC2GM092618, P50GM115305, U01HG006378, U19HL065962, R01HD074711.

The sequencing of 250,000 WGS individuals from BioVU®, including the 10,310 described here, has been funded by the Alliance for Genomic Discovery consisting of NashBio, Illumina and industry partners Amgen, AbbVie, AstraZeneca, Bayer, BMS, GSK, Merck, and Novo. DNA sequencing was performed at deCODE genetics using Illumina sequencing technology.

*Author contributions:*

RWC and YP are co–first authors. The authorship order reflects that the study was initiated by RWC, who was joined by YP in leading the project. RWC, YP, and AGB conceived the project. RWC performed statistical analyses and simulations with YP's input. CV performed CHIP calling. LYL, YP, TMM, KA, and CRC provided data from their respective studies with deep-targeted sequencing. RWC and YP drafted the initial manuscript. MRS made key conceptual contributions and critically revised the manuscript. All authors read and approved the final manuscript.

**Supplemental References**


1. Vlasschaert, C. *et al.* Clonal hematopoiesis of indeterminate potential is associated with acute kidney injury. *Nat. Med.* **30**, 810–817 (2024).
2. Crants, S. A. *et al.* Risk of clonal hematopoiesis of indeterminate potential after cancer radiation therapy. *medRxiv* 2024.09.27.24314321 (2024) doi:10.1101/2024.09.27.24314321.
3. Mack, T. *et al.* Germline genetics, disease, and exposure to medication influence longitudinal dynamics of clonal hematopoiesis. *Haematologica* (2024) doi:10.3324/haematol.2024.286513.
4. Amancherla, K. *et al.* Clonal hematopoiesis of indeterminate potential and outcomes after heart transplantation: A multicenter study. *Am. J. Transplant* **23**, 1256–1263 (2023).
